# Causal effect of the gut microbiota on the risk of psychiatric disorders and the mediating role of immunophenotypes

**DOI:** 10.1101/2024.07.29.24311128

**Authors:** Zhisheng Hong, Ao He, Guanglong Huang, Xiaofeng Chen, Xiaoyu Wang, Xiaoyang Li, Hailun Chen, Xinqi Zhao, Ying Xu, Yangheng Xu, Pei Ouyang, Hai Wang, Jiapeng Deng, Pengyu Chen, Xian Zhang, Songtao Qi, Yaomin Li

## Abstract

**Background:** Growing evidence indicates a significant correlation between the gut microbiota, immune system, and psychiatric disorders. Nevertheless, the impacts and interactions of the gut microbiota and immunophenotypes on psychiatric disorders remain unclear.

**Methods:** We utilized a bidirectional Mendelian randomization (MR) study to evaluate the causal associations among the gut microbiota, immunophenotypes, and psychiatric disorders, including attention-deficit/hyperactivity disorder (ADHD), major depressive disorder (MDD), posttraumatic stress disorder (PTSD), schizophrenia (SCZ), and Tourette’s syndrome (TS). The primary analysis was conducted using the inverse variance weighted (IVW) method, with several complementary sensitivity analyses being performed to ensure the reliability of the results.

**Results:** Our study reveals significant causal relationships between 22 immunophenotypes, 15 types of gut microbiota, and various psychiatric disorders. We further sought to ascertain whether immunophenotypes act as intermediaries in the pathway from gut microbiota to psychiatric disorders. In particular, three immunophenotypes were identified that mediate the causal effects of different gut microbiota on ADHD. Additionally, one immunophenotype was detected to mediate the causal effects of gut microbiota on PTSD.

**Conclusions:** Our study indicates that immunophenotypes partially mediate the pathway from the gut microbiota to psychiatric disorders.

## 1. Introduction

Psychiatric disorders, a kind of life-threatening and debilitating illness with heterogeneous etiology and complicated pathogenesis, incur a deadweight burden and have become a leading cause of disability worldwide [1]. Humanitarian emergencies, pandemics, and fierce competition have significantly increased the incidence of psychiatric disorders, affecting millions of people worldwide due to psychological stress [2]. Consequently, it is of the utmost importance to determine probable causal risk factors for various psychiatric disorders.

An increasing number of studies have reported that psychiatric disorders often overlap genetically and clinically, suggesting that they all share a similar underlying etiological mechanism [3–5]. Currently, the growing field of immuno-psychiatry acknowledges the pivotal role of the immune system in maintaining homeostasis and fortifying the resilience of central nervous system function [6]. Recent evidence indicates that immune dysregulation is involved in the pathophysiology of various psychiatric disorders [7–9]. Microglia, the brain’s macrophages, have been implicated in the pathogenesis of multiple psychiatric disorders, including depression, anxiety, and PTSD, through neuroinflammation [10–12]. Nevertheless, the causal associations between the immunophenotype and psychiatric disorders remain incomplete, unsystematic, and unclear.

Studies in humans and animals have reported microbial shifts associated with increased inflammation and alterations in host metabolism [13,14]. These microbes play a pivotal role in the maturation of the immune response, thereby contributing to homeostasis [15]. The microbiome profoundly influences peripheral immune pathways within the gut-brain communication axis, which regulate responses to neurogenesis [16], neuroinflammation [17], neurological injury [18], and neurodegeneration [19]. In addition, a substantial body of previous observational studies has demonstrated that there are discernible differences in the composition of the gut microbiota between healthy individuals and those diagnosed with various psychiatric disorders [20]. Nevertheless, the intricate nature of the gut microbiota makes it challenging to achieve a comprehensive and detailed understanding solely through observational studies.

A randomized controlled trial (RCT) is considered the most reliable method for establishing a causal relationship in the context of scientific research. However, the implementation of RCTs can be challenging or even unfeasible due to ethical restrictions. As an alternative, Mendelian randomization is an analytical approach that utilizes single nucleotide polymorphisms (SNPs) as an instrumental variable (IV) to investigate the causal relationship between an exposure or a risk factor and a clinically relevant outcome while minimizing the risk of bias and reverse causation [21]. In this study, we employed a systematic bidirectional two-sample MR design to comprehensively investigate the causal effects among the gut microbiome, immunophenotypes, and various psychiatric disorders. Subsequently, we sought to determine whether immunophenotypes act as mediators in the pathway from the gut microbiota to psychiatric disorders.

## 2. Methods

### 2.1 Study Design

This study comprises three principal phases. The initial phase involves an examination of the causal effects of 731 immunophenotypes on a range of major psychiatric disorders. This is followed by an investigation of the causal effects of 207 gut microbiota on psychiatric disorders. Finally, a mediation analysis was conducted to examine the role of immunophenotypes in the relationship between the gut microbiota and psychiatric disorders. Furthermore, a reverse MR analysis was conducted to circumvent the potential effects of reverse causality. The general design of our study is shown in Figure 1.

**Figure 1.**
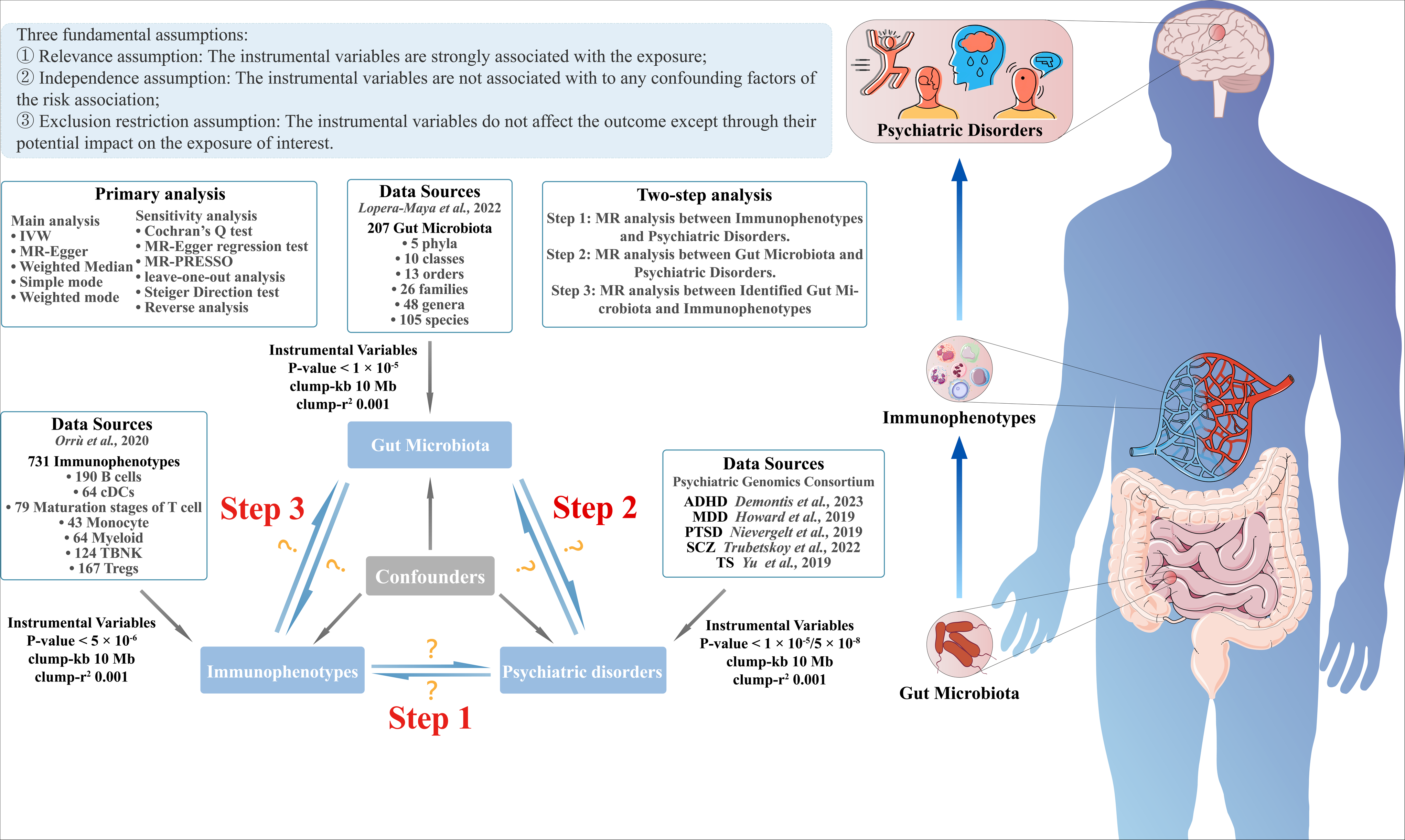
The general design of the study.

To determine the unbiased causal effects of exposure on outcomes, MR analysis must satisfy three fundamental assumptions: (I) relevance assumption: genetic variants are strongly correlated with exposures; (II) independence assumption: genetic variants are not associated with any confounding factors of the risk exposure-outcome association; (III) exclusion restriction assumption: genetic variants do not affect the outcome except through their potential impact on the exposure of interest [22,23]. This study followed Strengthening the Reporting of Observational Studies in Epidemiology (STROBE) Mendelian Randomization Reporting Guidelines [24].

### 2.2 Genome-wide Association Study Data Sources

Summary levels of the immunophenotype data were obtained from the latest GWAS [25]. The study analyzed a total of 731 immunophenotypes (3,757 Sardinians), comprising 118 absolute cell counts (AC), 389 median fluorescence intensities (MFI) reflecting surface antigen levels, 32 morphological parameters (MP), and 192 relative cell counts (RC). The GWAS data for the gut microbiome were generated from the Dutch Microbiome Project study, which involved a total of 207 gut microbiota (7,738 European individuals) to evaluate the impact of host genetics on the gut microbiota [26].

The GWAS summary data for ADHD [27], MDD [28], PTSD [29], SCZ [30], and TS [31] were derived from the Psychiatric Genomics Consortium (PGC). All cohorts were case-control studies, and cases met the criteria defined in the International Statistical Classification of Diseases and Related Health Problems-10th Revision (ICD-10) diagnosis code or medication prescription.

All participants were of European ancestry. The original publications contain detailed diagnostic criteria and methods used to recruit participants for these GWASs. There was no significant overlap between the GWAS datasets. The characteristics of the selected GWAS data are listed in Table S1.

### 2.3 Instrumental Variables Selection

First, SNPs significantly associated with the immunophenotypes (P < 5×10^−6^) and the gut microbiota (P < 1×10^−5^) were selected [32,33]. To guarantee the premise of independence, IVs were pruned with linkage disequilibrium (LD) r^2^ > 0.001 in the 1000 Genomes European data within 10 Mb windows [34]. To compensate for missing SNPs, those with strong linkage disequilibrium (r^2^ > 0.8) were utilized [35]. The proportion of variance in the explained phenotype (R^2^) was calculated to indicate the power of MR studies [36]. The F-statistic of each IV was calculated using the formula [R^2^/(1-R^2^)] × [(N-K-1)/K] [37]. To prevent weak instrument bias from influencing the results, any IV with an F-statistic less than 10 was removed from consideration. The summary statistics were harmonized, and any mismatched strands or palindromic SNPs were removed [38]. Additionally, the summary statistics were aligned to ensure that each genetic variant was associated with the same effect allele. The research findings are deemed reliable due to the meticulous selection of IVs.

### 2.4 Primary Analysis

Two-sample MR analysis was conducted using the multiplicative random effects inverse variance weighted approach to evaluate the causal effects of the gut microbiota and immunophenotype on psychiatric disorders [39]. To evaluate the robustness of the primary estimates, the MR-Egger and Weighted Median methods were employed to corroborate the IVW results [40,41]. The coherence in the orientation of the odds ratio (OR) for IVW, MR-Egger, and weighted median was deemed essential for the robustness of the findings. The false discovery rate (FDR) was utilized to adjust for multiple tests, and a significant causal relationship was established using an FDR-adjusted threshold of P < 0.05 [42]. A P-value of less than 0.05, but above the FDR threshold, was considered indicative of a nominal association.

### 2.5 Mediation Analysis

Following the bidirectional analysis, the significant causal associations between the gut microbiota or immunophenotype, and psychiatric disorders were included in the two-step mediation analysis. Subsequently, the mediation proportions were calculated according to the following formula: (ß1 × ß2) / ß, where ß1 indicates the impact of the gut microbiota on mediators, ß2 indicates the impact of mediators on psychiatric disorders, and ß indicates the total impact of the gut microbiota on psychiatric disorders [43]. Standard errors and confidence intervals (CIs) were calculated using delta methods [44]. Effect estimates are reported as beta values (ß) for the continuous outcome and ORs for the binary outcome.

### 2.6 Sensitivity Analysis

Cochran’s Q statistic was used to assess the heterogeneity across the individual causal effects [45]. A leave-one-out analysis was conducted to detect outlier instrumental variables [46]. MR-Egger regression and MR pleiotropy residual sum and outlier (MR-PRESSO) analyses were performed to evaluate potential horizontal pleiotropy [47,48]. Outliers identified by the MR-PRESSO outlier test were removed step-by-step to reduce the effect of horizontal pleiotropy, and a corrected casual result was recalculated. Furthermore, the Steiger test was employed to negate the potential bias resulting from reverse causality [49].

### 2.7 Reverse Analysis

To assess the causal effects of psychiatric disorders on the gut microbiota and immunophenotypes, a backward design was employed, with psychiatric disorders serving as the exposures and the gut microbiota or immunophenotypes serving as the outcomes. SNPs significantly associated with ADHD, MDD, and SCZ at the genome-wide significance level (P < 5×10^−8^) were selected, while a higher cutoff (P < 1×10^−5^) was used for PTSD and TS.

All analyses were conducted using R statistical software version 4.3.1 (accessed on October 18, 2023, https://www.r-project.org/). MR analysis was performed using the R-based packages “TwoSampleMR” (v.0.5.9) [50] and “MR_PRESSO” [51].

## 3. Results

### 3.1 Instrumental Variables Selection

After performing clumping and linkage disequilibrium pruning, we identified 10,613 SNPs associated with immunophenotypes and 1,962 SNPs related to the gut microbiota. For the reverse analysis, we selected genetic instruments associated with psychiatric disorders, with SNPs ranging from 26 to 155. All instrumental variables exhibited F-statistics exceeding 10, thereby demonstrating a robust association with exposure phenotypes. Further details on the specific genetic instrumental variables used to infer causal effects are provided in Table S2.

### 3.2 Causal Effects of Immunophenotypes on Psychiatric Disorders

As shown in Figure 2 and Table S3A, we identified 275 potential causal associations between immunophenotypes and five psychiatric disorders. Multiple tests identified 22 significant causal associations, as shown in Figure 3. The results of the subsequent sensitivity analysis were robust to the causal effect analysis, as shown in Tables S3B and S3C.

**Figure 2.**
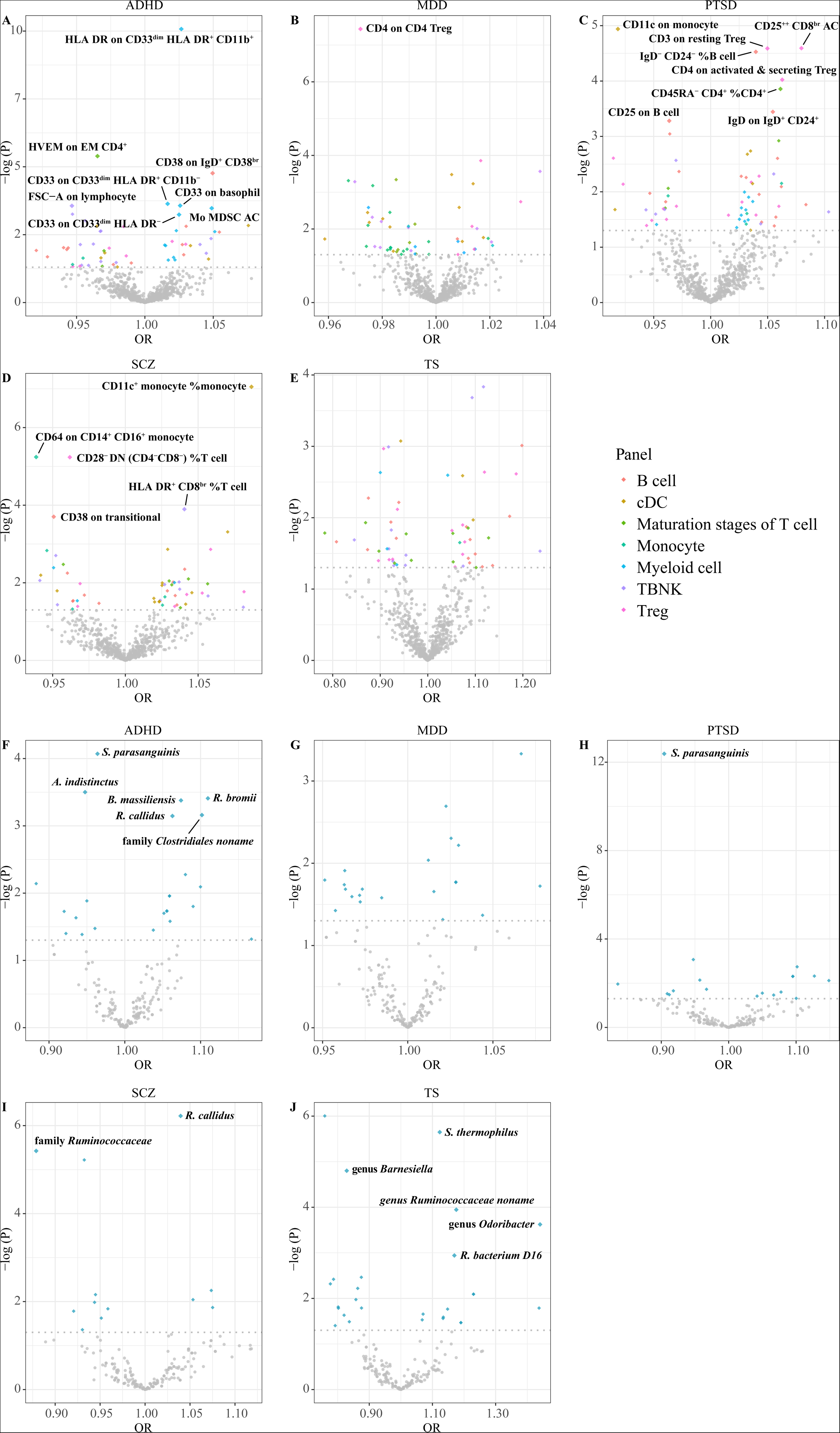
Volcano plots showing the effect estimates for each exposure to psychiatric disorders. A-E demonstrate the causal effect of immunophenotypes on psychiatric disorders. F-J demonstrate the causal effect of the gut microbiota on psychiatric disorders.

**Figure 3.**
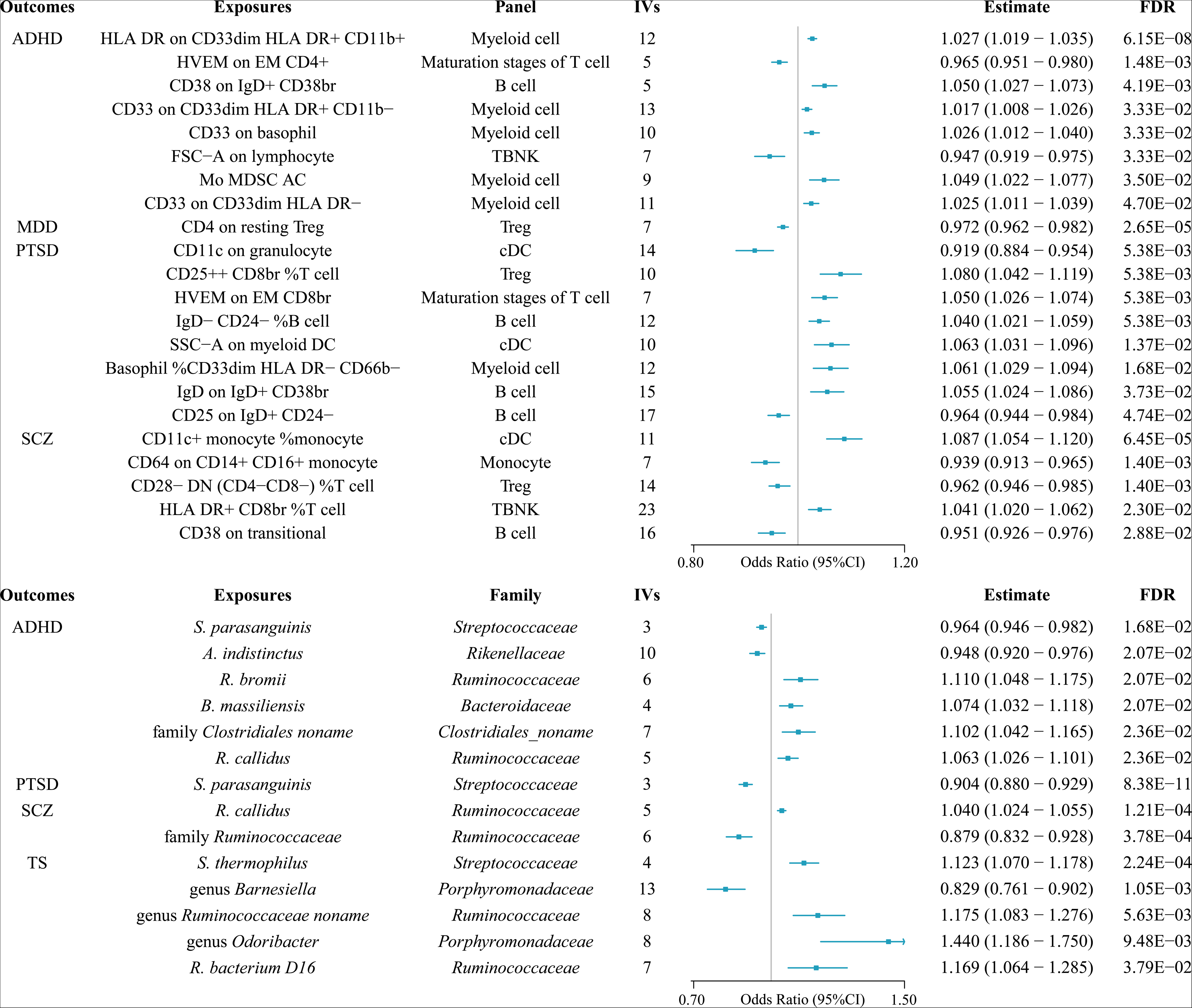
Mendelian randomization results of causal effects between immunophenotypes, the gut microbiota, and psychiatric disorders.

We identified 54 potential causal associations between immunophenotypes and ADHD. After adjusting for multiple tests, we investigated five immunophenotypes that contribute causally to the risk of ADHD from the myeloid cell panel, namely, the level of HLA DR on CD33^dim^ HLA DR^+^ CD11b^+^ (OR = 1.027, 95% CI 1.019 - 1.035, P = 8.41×10^−11^), the level of CD33 on CD33^dim^ HLA DR^+^ CD11b^−^ (OR = 1.017, 95% 1.008 - 1.026, P = 2.31×10^−4^), the level of CD33 on basophils (OR = 1.026, 95% CI 1.012 - 1.040, P = 2.69×10^−4^), the absolute level of monocytic myeloid-derived suppressor cells (Mo MDSCs) (OR = 1.049, 95% CI 1.022 - 1.077, P = 3.35×10^−4^), and the level of CD33 on CD33^dim^ HLA DR^−^ (OR = 1.025, 95% CI 1.011 - 1.039, P = 5.78×10^−4^). Furthermore, the genetic increase in the level of herpes virus entry mediator (HVEM) on effector memory (EM) CD4^+^ T cells (OR = 0.965, 95% CI 0.951 - 0.980, P = 4.04×10^−6^) was potentially related to a decreased risk of ADHD. Conversely, the level of CD38 on IgD^+^ CD38^br^ (OR = 1.050, 95% CI 1.027 - 1.073, P = 1.72×10^−5^) increased the incidence of ADHD. Interestingly, our findings indicate a protective effect of elevated lymphocyte FSC-A parameters (OR = 0.947, 95% CI 0.919 - 0.975, P = 2.74×10^−4^) on the risk of ADHD from a morphological perspective.

However, in leave-one-out analyses, we found that rs3865444 would significantly drive the causal inference of the level of CD33 on CD33^dim^ HLA DR^+^ CD11b^−^, while rs190188611 would significantly drive the causal inference of lymphocyte FSC-A parameters, respectively. Consequently, both phenotypes were excluded from subsequent mediation analyses, given the robustness of the conclusions.

We identified 52 potential causal associations between immunophenotypes and MDD. After adjusting for multiple tests, we identified the level of CD4 on resting Tregs (OR = 0.972, 95% CI 0.962 - 0.982, P = 3.66×10^−8^) as a significant protective factor against the risk of MDD.

We identified 63 potential causal associations between immunophenotypes and PTSD. After adjustment for multiple tests, genetic prediction of two immunophenotypes was associated with a decreased risk of PTSD: the level of CD11c on granulocytes (OR = 0.919, 95% CI 0.884 - 0.954, P = 1.15×10^−5^) and the level of CD25 on IgD^+^ CD24^−^ (OR = 0.964, 95% CI 0.944 - 0.984, P = 5.24×10^−5^). Furthermore, six phenotypes were identified as being associated with an increased risk of PTSD: the relative percentage of CD25^++^ CD8^br^ among T cells (OR = 1.080, 95% CI 1.042 - 1.119, P = 2.55×10^−5^), the level of HVEM on EM CD8^br^ (OR = 1.050, 95% CI 1.026 - 1.074, P = 2.58×10^−5^), the relative percentage of IgD^−^ CD24^−^ among B cells (OR = 1.040, 95% CI 1.021 - 1.059, P = 2.97×10^−5^), the myeloid DC SSC-A parameter (OR = 1.063, 95% CI 1.031 - 1.096, P = 9.46×10^−5^), the relative percentage of basophil among CD33^dim^ HLA DR^−^ CD66b^−^ (OR = 1.061, 95% CI 1.029 - 1.094, P = 1.39×10^−4^), and the level of IgD on IgD^+^ CD38^br^ (OR = 1.055, 95% CI 1.024 - 1.086, P = 3.60×10^−4^).

However, in leave-one-out analyses, the causal inference of the myeloid DC SSC-A parameter was significantly driven by rs189299852. Consequently, this phenotype was excluded from subsequent mediation analyses.

We identified 53 potential causal associations between immunophenotype and SCZ. After adjusting for multiple tests, the relative percentage of CD11c^+^ monocytes among monocytes (OR = 1.087, 95% CI 1.054 - 1.120, P = 8.90×10^−8^), and the relative percentage of HLA DR^+^ CD8^br^ among T cells (OR = 1.041, 95% CI 1.020 - 1.062, P = 1.27×10^−4^) were identified as risk factors. Furthermore, the level of CD64 on CD14^+^ CD16^+^ monocytes (OR = 0.939, 95% CI 0.913 - 0.965, P = 5.74×10^−6^), the relative percentage of CD28^−^ DN (CD4^−^CD8^−^) among T cells (OR = 0.962, 95% CI 0.946 - 0.985, P = 5.81×10^−6^), and the level of CD38 on transitional cells (OR = 0.951, 95% CI 0.926 - 0.976, P = 1.99×10^−4^) were identified as protective factors.

We identified 53 potential causal associations between immunophenotype and TS. Nevertheless, following the application of multiple test correction, no immunophenotype was found to be significantly associated with TS.

### 3.3 Causal Effects of the Gut Microbiota on Psychiatric Disorders

As shown in Figure 2 and Table S3D, we identified 66 potential causal associations between the gut microbiota and five psychiatric disorders. Multiple tests identified 14 significant causal associations, as shown in Figure 3. The results of the subsequent sensitivity analysis were robust to the causal effect analysis, as shown in Tables S3E and S3F.

A total of 19 nominal causal effects of the gut microbiota on ADHD were identified. After adjustment for the P-value, the MR analysis indicated that the genetic predictions of *Streptococcus parasanguinis* (OR = 0.964, 95% CI 0.946 - 0.982, P = 8.49×10^−5^) and *Alistipes indistinctus* (OR = 0.948, 95% CI 0.920 - 0.976, P = 3.16×10^−4^) were associated with a decreased risk of ADHD. Conversely, *Ruminococcus bromii* (OR = 1.110, 95% CI 1.048 - 1.175, P = 3.90×10^−4^) and *Ruminococcus callidus* (OR = 1.063, 95% CI 1.026 - 1.101, P = 7.14×10^−4^) significantly increased the incidence of ADHD. Furthermore, *Bacteroides massiliensis* (OR = 1.074, 95% CI 1.032 - 1.118, P = 4.19×10^−4^) was potentially related to a greater risk of ADHD. Interestingly, an unnamed family from the order *Clostridiales* (OR = 1.102, 95% CI 1.042 - 1.165, P = 6.91×10^−4^) was identified as a risk factor for ADHD.

A total of eight nominal causal effects of the gut microbiota on MDD were identified. After adjustment for the P-value, no gut microbiota was found to be significantly associated with MDD.

Thirteen nominal causal effects of the gut microbiota on PTSD were identified. After adjustment for the P-value, *S. parasanguinis* (OR = 0.904, 95% CI 0.880 - 0.929, P = 4.17×10^−13^) was found to be significantly associated with a decreased risk of PTSD, similar to that observed for ADHD.

A total of five nominal causal effects of the gut microbiota on SCZ were identified. After adjustment for the P-value, genetic prediction of the family *Ruminococcaceae* (OR = 0.879, 95% CI 0.832 - 0.928, P = 3.76×10^−6^) was associated with a decreased risk of SCZ. Notably, *R. callidus* (OR = 1.040, 95% CI 1.024 - 1.055, P = 6.02×10^−7^), from the family *Ruminococcaceae*, was determined to be significantly associated with increased susceptibility to SCZ, similar to that observed for ADHD.

A total of 21 nominal causal effects of the gut microbiota on TS were identified. After adjustment for the P-value, the genetic prediction of four gut microbiota (including two genera and two species) was found to be associated with an increased risk of TS. These included *Streptococcus thermophilus* (OR = 1.123, 95% CI 1.070 - 1.178, P = 2.25×10^−6^), *Ruminococcaceae bacterium D16* (OR = 1.169, 95% CI 1.064 - 1.285, P = 1.14×10^−3^), genus *Odoribacter* (OR = 1.440, 95% CI 1.186 - 1.750, P = 2.38×10^−4^), and an unnamed genus from the family *Ruminococcaceae* (OR = 1.175, 95% CI 1.083 - 1.276, P = 1.13×10^−4^). Nevertheless, the genus *Barnesiella* (OR = 0.829, 95% CI 0.761 - 0.902, P = 1.58×10^−5^), which belongs to the same family (family *Porphyromonadaceae*) as the genus *Odoribacter*, was demonstrated to exert a protective effect against TS.

### 3.4 Mediation Analysis

As shown in Figure 4 and Table S4, the gut microbiota and immunophenotype were found to exert causal effects on psychiatric disorders, with the immunophenotype serving as a partial mediator in the pathway from the gut microbiota to ADHD and PTSD. However, in leave-one-out analyses, the causal inferences of *R. bromii* on the level of CD33 on basophils and the level of CD33 on CD33^dim^ HLA DR^−^ were significantly driven by rs12041621 and rs1884673. Consequently, these phenotypes were excluded from mediation analysis to prevent the introduction of statistical bias due to the influence of a single SNP.

**Figure 4.**
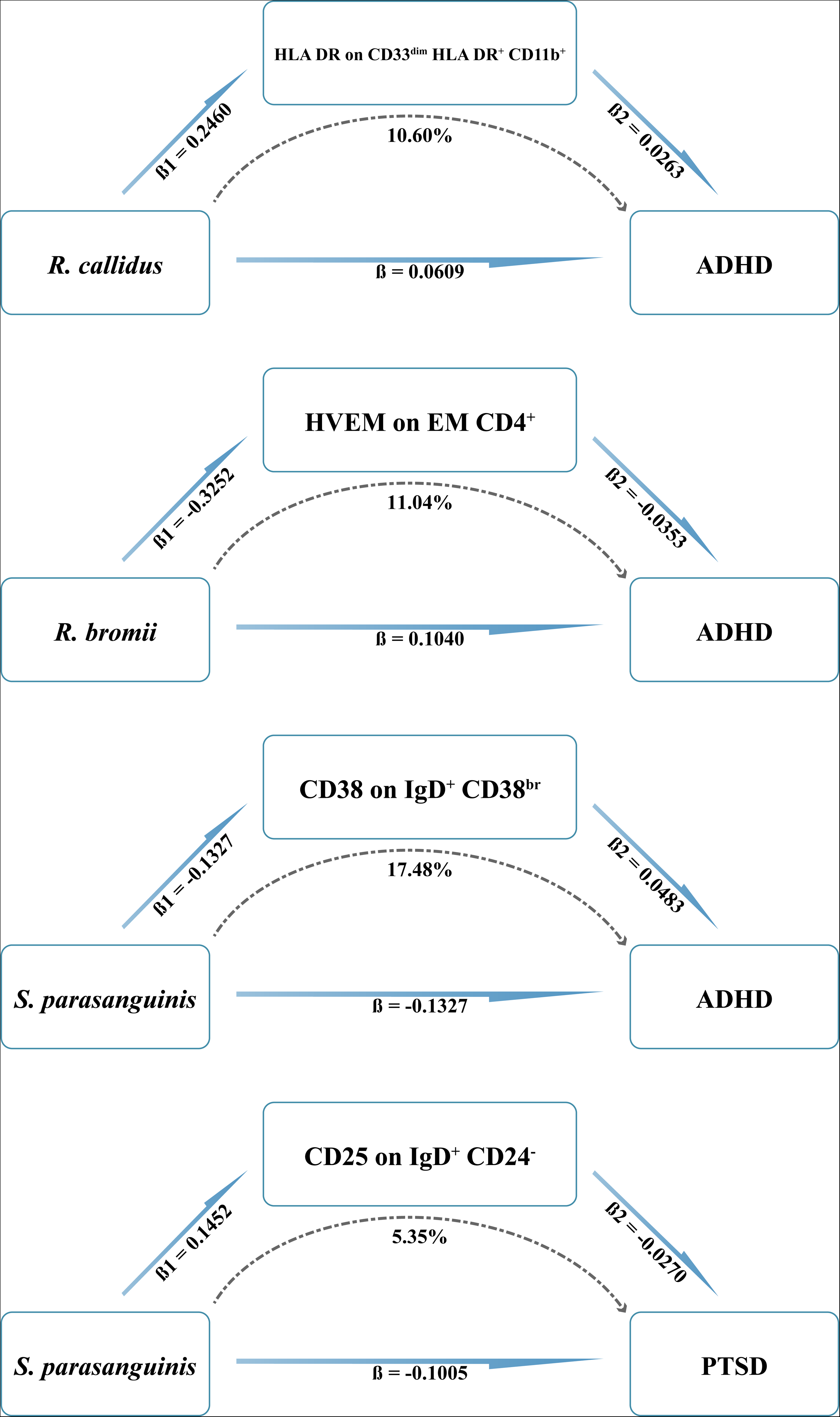
Immunophenotypes mediate the causal pathways from the gut microbiota to ADHD and PTSD.

Significant associations were identified between the genetic prediction of *R. bromii* and a decreased level of HVEM on EM CD4^+^ (ß = −0.325, 95% CI −0.557 - −0.093, P = 5.95×10^−3^), mediated 11.04% of the causal effect of *R. bromii* on ADHD. Furthermore, we observed suggestive evidence indicating that an elevated level of *R. callidus* was associated with a significantly greater level of HLA DR on CD33^dim^ HLA DR^+^ CD11b^+^ (ß = 0.246, 95% CI 0.173 - 0.319, P = 5.14×10^−11^), which mediated 10.60% of the causal effect of *R. callidus* on ADHD. Notably, a potential association was identified between the genetic prediction of *S. parasanguinis* and a decreased level of CD38 on IgD^+^ CD38^br^ (ß = −0.132, 95% CI −0.211 - −0.054, P = 9.20×10^−4^), which accounted for 17.48% of the causal effect of *S. parasanguinis* on ADHD. Conversely, the genetic prediction of *S. parasanguinis* was an increase in the level of CD25 on IgD^+^ CD24^−^ (ß = 0.145, 95% CI 0.067 - 0.224, P = 3.04×10^−4^), which contributed to 5.35% of the causal effect of *S. parasanguinis* on PTSD.

### 3.5 Reverse Analysis

According to the backward analysis, as shown in Tables S5A and S5B, the onset of ADHD significantly decreased the relative abundance of *Coprococcus catus* (ß = −0.222, 95% CI −0.311 - −0.133, P = 1.04×10^−6^). Furthermore, we sought to ascertain whether the previously identified significant causality exhibited reverse interference. As shown in Tables S5C, S5D and S5E, our findings indicate that reverse causality was not a potential factor in these instances.

### 3.6 Sensitivity Analysis

The Cochran’s Q test indicated the absence of significant heterogeneity. No evidence of horizontal pleiotropy was identified, as the intercept of MR-Egger did not significantly deviate from zero. Additionally, no potential instrumental outliers were detected at the nominal significance level of 0.05 by MR-PRESSO analysis. The leave-one-out results indicate that no single instrumental variable drove the remaining causal effects. These results serve to confirm the validity of our analysis and demonstrate its robustness in avoiding unintentional errors.

## 4. Discussion

This study presents, for the first time, multiple novel potential associations between genetically predicted gut microbiota or immunophenotypes and psychiatric disorders using bidirectional MR analysis with mediation analysis. In addition, we identified multiple potential pathways of immunophenotype-mediated causality between the gut microbiota and psychiatric disorders. This provides a new direction for understanding how the gut-brain axis may influence psychiatric disorders.

ADHD is a neurodevelopmental disorder that persists, characterized by inattention, hyperactivity, and impulsivity [52]. The prevalence of ADHD varies across populations and age groups, and a meta-analysis suggested that ADHD affects 5% of children and adolescents and 2.5% of adults worldwide [53]. This study demonstrated that *R. callidus* is associated with an increased risk of ADHD, which is mediated by elevated expression of HLA DR on the CD33^dim^ HLA DR^+^ CD11b^+^ surface. HLA DR gene region has been implicated in several psychiatric disorders, including schizophrenia, bipolar disorder, and autism spectrum disorders [54]. Nevertheless, current research has been unable to identify a correlation between ADHD and HLA. Further studies are necessary to confirm this hypothesis. Moreover, our findings also indicate that *R. bromii* may increase the risk of ADHD by reducing the expression of HVEM on EM CD4^+^, which is regarded as a protective factor for ADHD. However, few previous studies have suggested this, which may represent a novel perspective that diverges from the previous belief that *R. bromii* is a probiotic [55]. Notably, HVEM plays a role in the maintenance of T-cell immune homeostasis [56]. This phenomenon may be attributed to the diminished coinhibitory effect of HVEM, which stimulates effector memory T cells to secrete copious quantities of cytokines (e.g., IL-17 and IFN-γ) [57,58]. These cytokines trigger neuroinflammation, a process that could contribute to the etiology of ADHD.

This study demonstrated that elevated levels of various myeloid cell phenotypes, particularly high surface expression of CD33, are significantly and positively associated with the risk of developing ADHD. CD33 is a molecule of the Siglec family that is mainly expressed in myeloid cells, including macrophages and dendritic cells, and is involved in regulating cellular activation, differentiation, and inflammatory responses [59]. In addition, microglia, a vital subset of immune cells within the central nervous system, also express CD33 molecules [60]. Synaptic pruning mediated by microglia involves the elimination of excessive or weak synaptic connections, thereby strengthening the more important and efficient neural pathways, which, if their activities are abnormal, can affect neural circuitry development, leading to ADHD-related symptoms [61,62]. Furthermore, under conditions of focal brain injury or neuroinflammation, Mo MDSCs can infiltrate the brain and inhibit neuronal inflammation by suppressing microglia activation, which is triggered by factors like IFN-γ, GM-CSF, and TNF [63,64].

Surprisingly, this study revealed that ADHD could decrease the abundance of *C. catus*. Previous studies have shown that the Mediterranean diet can increase the abundance of C. catus [65]. It was postulated that patients with ADHD may experience appetite suppression as a side effect of medication or as a consequence of abnormal eating habits resulting from mood changes associated with the onset of ADHD. Consequently, this could result in a reduction in the abundance of *C. catus*. Furthermore, a reduction in the abundance of *C. catus* may result in a decrease in butyrate production, which plays an important anti-inflammatory role in the central nervous system [66,67]. This, in turn, may exacerbate ADHD symptoms. Consequently, fluctuations in the prevalence of *C. catus* can be employed to inform timely modification of the dietary regimen of ADHD patients, thereby preventing exacerbation of the disease by establishing a diverse and healthy gut microbiota.

PTSD is a debilitating mental disorder with a lifetime prevalence of nearly 8% in the general population [68]. Those with PTSD experience a constant state of hyperarousal and fear, which may be associated with abnormal gene expression in the brain and peripheral blood cells [69]. Furthermore, the coaggregation of PTSD and ADHD within families implies the presence of shared familial risk factors for these disorders [70]. In this investigation, *S. parasanguinis* was identified as a co-protective element against ADHD and PTSD. This effect was found to be mediated by CD38 on IgD^+^ CD38^br^ and CD25 on IgD^+^ CD24^−^. *S. parasanguinis* is a gram-positive coccus that typically forms a symbiotic relationship with the human body and has the capacity to suppress the growth of pathogenic bacteria through the secretion of antibacterial substances [71,72]. Notably, *S. parasanguinis* is known to colonize the gastrointestinal tract and stimulate gene expression in Tregs, resulting in the enhancement of anti-inflammatory cytokine secretion, thus indicating its immunomodulatory capabilities [73]. Given the established associations between ADHD and oxidative stress, as well as neuroinflammation, it can be postulated that *S. parasanguinis* might exert an immunosuppressive effect through the downregulation of CD38, which may play a role in promoting neurodegeneration and inflammatory harm [74,75]. Equally significant is the role of CD25 in suppressing the activation of self-reactive T cells through mechanisms that are independent of cytokines and reliant on cell contact to promote immune tolerance [76,77]. *S. parasanguinis* has the potential to alleviate chronic low-grade inflammation induced by prolonged stress through the upregulation of CD25, which may contribute to immune regulation, thereby exerting a protective effect on PTSD. Furthermore, the immunophenotypes closely linked to the development of PTSD predominantly consist of B lymphocytes. These cells may enter the prefrontal cortex, insula, amygdala, hippocampus, and other brain regions either by crossing the blood-brain barrier due to the peripheral release of proinflammatory cytokines or by directly infiltrating the central nervous system [78]. This process can intensify the manifestations of fear and anxiety. Further investigation is required to elucidate the potential mechanisms of targeted B-cell intervention for the alleviation or management of PTSD.

SCZ is a severe mental illness characterized by positive symptoms and cognitive impairment [79]. Growing evidence indicates that the gut microbiota may influence social behavior, emotion, and cognition via the gut-brain axis, with the potential to impact the development of SCZ [80]. Our findings suggest that the increased abundance of *Ruminococcus* may be a protective factor for SCZ. *Ruminococcus* species are believed to produce butyrate, which can enhance antioxidant capacity and regulate inflammatory mediators. Surprisingly, we also found that the increased abundance of R. callidus may be a risk factor for SCZ, similar to that observed in ADHD patients. Furthermore, our study demonstrated a significant correlation between reduced levels of CD38 in transitional cells and increased susceptibility to SCZ. This finding is substantiated by animal experiments, which underscore the crucial involvement of CD38 in neurodevelopmental mechanisms such as neuropeptide release and social cognition [81]. Moreover, immunophenotypes that include CD8 molecules may increase susceptibility to SCZ, resembling MDD, which is characterized by a reduced CD4/CD8 ratio in acute psychosis episodes [82]. The underlying mechanism remains ambiguous, with speculation suggesting a potential association with systemic inflammation triggered by adaptive immunity. In addition, various subtypes of monocytes, including the relative percentage of CD11c+ monocytes among monocytes and the level of CD64 on CD14+ CD16+ monocytes, have been shown to exhibit both detrimental and beneficial effects on SCZ. This duality in effects may be attributed to the heterogeneity of monocytes circulating in vivo, which display a combination of proinflammatory and anti-inflammatory traits [83].

MDD is a pervasive mental disorder characterized by persistent low mood, sleep disturbances, cognitive decline, and other symptoms [84]. There is a plethora of evidence indicating that neuroinflammation can precipitate cellular immune disorders and increase the risk of psychiatric disorders, and that Tregs play a pivotal role in regulating the immune response [85]. MDD typically involves stress activation by the immune system, leading to the release of stressor-induced circulating proinflammatory cytokines [86,87]. Consequently, CD4^+^CD25^+^ Tregs with immunomodulatory functions may significantly reduce anxiety-like behaviors by inhibiting proinflammatory cytokines such as Th1, Th2, and Th17 cells, thereby alleviating MDD [88]. Importantly, a significant increase in Tregs was previously observed during antidepressant treatment [89], which strongly supports our conclusion. In light of the aforementioned studies, we hypothesized that the level of CD4 on resting Tregs subsets plays an important protective role in the pathogenesis of MDD by regulating neuroinflammation and reducing oxidative stress.

TS is a neurological disorder characterized by repetitive and involuntary movements and vocalizations, and can be divided into motor tics and vocal tics. These symptoms often begin in childhood. A significant body of research has demonstrated that the gut-brain axis plays a pivotal role in the pathophysiology of TS [90]. Fecal microbiota transplantation (FMT) has been identified as a potential treatment for various neurological and psychiatric disorders, including TS, by regulating the microbiota ecology to maintain cytokine balance in the gut lymphoid tissue and reduce the release of proinflammatory factors [91]. *S. thermophilus* is a safe probiotic with anti-inflammatory, anti-cancer, and antioxidant properties [92]. However, our results showed that a high abundance of *S. thermophilus* was a risk factor for TS, in contrast to previous studies. Given the paucity of studies investigating the relationship between *S. thermophilus* and TS, further research on its mechanism is warranted. In addition, our study demonstrated that the increased abundance of *R. noname*, *G. odoribacter*, and *R. bacterium D16* may be risk factors for TS. This may be because *R. noname* and *R. bacterium D16* both belong to the *Ruminococcaceae* family, and the variation in *R. UCG-004* is positively correlated with the anxiety-like behavior of individuals with ADHD. Importantly, the comorbidity rate of ADHD and TS was 50%, which led us to infer that *G. odoribacter* and *R. bacterium D16* may be common risk factors for both [93,94]. In fact, according to a study by Wang et al., *G. odoribacter* may interfere with the dopamine metabolic pathway to cause TS pathogenesis, which is consistent with our conclusion [95]. Finally, we found that increased abundance of *G. barnesiella* reduced the risk of TS, possibly because *G. barnesiella* is a natural gut microbe involved in competitive inhibition of pathogenic bacteria and immune regulation, improving cognitive function and thereby ameliorating TS-related symptoms [96,97].

There are several limitations to consider in this study. The current GWAS database constraints hinder the analysis of data from other ethnic groups, and caution is advised when interpreting our results in different populations. Moreover, given the inherent diversity among patients with psychiatric disorders, future research could delve into these subgroups in more detail. Finally, further prospective controlled trials may be necessary to investigate the mechanisms underlying the potential roles of recently discovered gut microbiota and immunophenotypes in psychiatric disorders.

## 5. Conclusion

In summary, our study comprehensively assessed the significant causal relationships between 22 immunophenotypes, 15 gut microbiota, and five psychiatric disorders. Furthermore, four potential causal pathways between gut microbiota, immunophenotypes, and psychiatric disorders were identified. This highlights the intricate pattern of interactions between the gut microbiota, immune system, and psychiatric disorders. These findings provide valuable insights for risk assessment and potential treatment strategies. Consequently, a more comprehensive understanding of the underlying mechanisms governing these causal relationships could facilitate the early identification, intervention, and prevention of psychiatric disorders.

## Supporting information

STROBE-MR checklist

## Data Availability

The datasets analyzed in this study are publicly available summary statistics. The data can be obtained through cited papers and websites.

https://www.ebi.ac.uk/gwas/

## List of abbreviations

MR: Mendelian randomization
ADHD: Attention-deficit/hyperactivity disorder
MDD: Major depressive disorder
PTSD: Posttraumatic stress disorder
SCZ: Schizophrenia
TS: Tourette’s syndrome
IVW: Inverse variance weighted
RCT: Randomized controlled trial
SNP: Single nucleotide polymorphism
IV: Instrumental variable
GWAS: Genome-wide association studies
LD: Linkage disequilibrium
OR: Odds ratio
FDR: False discovery rate
CI: Confidence intervals
MR-PRESSO: MR pleiotropy residual sum and outlier analysis
MDSC: myeloid-derived suppressor cells
HVEM: Herpes virus entry mediator

## Supplementary Information

Additional file 1: Supplementary Tables.xlsx.

Additional file 2: STROBE-MR checklist.docx.

## Acknowledgement

The authors would like to express their gratitude to the investigators of the original studies for sharing the GWAS summary statistics.

## Declarations

### Ethics approval and consent to participate

All data used in this work are publicly available from studies with relevant participant consent and ethical approval.

### Consent for publication

Not applicable.

### Authors’ contributions

Z.H., A.H., and G.H. conceived and designed the project. Z.H., and A.H. drafted the manuscript. X.C., X.W., X.L., H.C., and X.Z. investigated and collected the data. Z.H., Y.X.1, Y.X.2, and P.O. supervised and analyzed the data. A.H., G.H., and H.W. conducted validation. J.D., and P.C. visualized the results. Y.L., S.Q., and X.Z. revised the manuscript. All authors carefully revised the manuscript and approved the version submitted.

### Code availability

All R scripts applied in this study are available from the authors upon reasonable request.

## References

[1] Collins PY, Patel V, Joestl SS, March D, Insel TR, Daar AS, et al. Grand challenges in global mental health. Nature 2011;475:27–30. 10.1038/475027a.

[2] Taquet M, Luciano S, Geddes JR, Harrison PJ. Bidirectional associations between COVID-19 and psychiatric disorder: retrospective cohort studies of 62 354 COVID-19 cases in the USA. Lancet Psychiatry 2021;8:130–40. 10.1016/S2215-0366(20)30462-4.

[3] Momen NC, Plana-Ripoll O, Agerbo E, Benros ME, Børglum AD, Christensen MK, et al. Association between Mental Disorders and Subsequent Medical Conditions. N Engl J Med 2020;382:1721–31. 10.1056/NEJMoa1915784.

[4] Plana-Ripoll O, Pedersen CB, Holtz Y, Benros ME, Dalsgaard S, De Jonge P, et al. Exploring Comorbidity Within Mental Disorders Among a Danish National Population. JAMA Psychiatry 2019;76:259. 10.1001/jamapsychiatry.2018.3658.

[5] Kessler RC, Avenevoli S, McLaughlin KA, Green JG, Lakoma MD, Petukhova M, et al. Lifetime co-morbidity of DSM-IV disorders in the US National Comorbidity Survey Replication Adolescent Supplement (NCS-A). Psychol Med 2012;42:1997–2010. 10.1017/S0033291712000025.

[6] Pape K, Tamouza R, Leboyer M, Zipp F. Immunoneuropsychiatry — novel perspectives on brain disorders. Nat Rev Neurol 2019;15:317–28. 10.1038/s41582-019-0174-4.

[7] Sewell MDE, Jiménez-Sánchez L, Shen X, Edmondson-Stait AJ, Green C, Adams MJ, et al. Associations between major psychiatric disorder polygenic risk scores and blood-based markers in UK biobank. Brain Behav Immun 2021;97:32–41. 10.1016/j.bbi.2021.06.002.

[8] Kwon S, Cheon SY. Influence of the inflammasome complex on psychiatric disorders: clinical and preclinical studies. Expert Opin Ther Targets 2021;25:897–907. 10.1080/14728222.2021.2005027.

[9] Zhang R, Song J, Isgren A, Jakobsson J, Blennow K, Sellgren CM, et al. Genome-wide study of immune biomarkers in cerebrospinal fluid and serum from patients with bipolar disorder and controls. Transl Psychiatry 2020;10:58. 10.1038/s41398-020-0737-6.

[10] Mondelli V, Vernon AC, Turkheimer F, Dazzan P, Pariante CM. Brain microglia in psychiatric disorders. Lancet Psychiatry 2017;4:563–72. 10.1016/S2215-0366(17)30101-3.

[11] Smith KL, Kassem MS, Clarke DJ, Kuligowski MP, Bedoya-Pérez MA, Todd SM, et al. Microglial cell hyper-ramification and neuronal dendritic spine loss in the hippocampus and medial prefrontal cortex in a mouse model of PTSD. Brain Behav Immun 2019;80:889–99. 10.1016/j.bbi.2019.05.042.

[12] Kreisel T, Frank MG, Licht T, Reshef R, Ben-Menachem-Zidon O, Baratta MV, et al. Dynamic microglial alterations underlie stress-induced depressive-like behavior and suppressed neurogenesis. Mol Psychiatry 2014;19:699–709. 10.1038/mp.2013.155.

[13] Westfall S, Caracci F, Zhao D, Wu Q, Frolinger T, Simon J, et al. Microbiota metabolites modulate the T helper 17 to regulatory T cell (Th17/Treg) imbalance promoting resilience to stress-induced anxiety- and depressive-like behaviors. Brain Behav Immun 2021;91:350–68. 10.1016/j.bbi.2020.10.013.

[14] Ling Z, Liu X, Cheng Y, Yan X, Wu S. Gut microbiota and aging. Crit Rev Food Sci Nutr 2022;62:3509–34. 10.1080/10408398.2020.1867054.

[15] O’Hara AM, Shanahan F. The gut flora as a forgotten organ. EMBO Rep 2006;7:688–93. 10.1038/sj.embor.7400731.

[16] Humann J, Mann B, Gao G, Moresco P, Ramahi J, Loh LN, et al. Bacterial Peptidoglycan Traverses the Placenta to Induce Fetal Neuroproliferation and Aberrant Postnatal Behavior. Cell Host Microbe 2016;19:388–99. 10.1016/j.chom.2016.02.009.

[17] Rea K, Dinan TG, Cryan JF. The microbiome: A key regulator of stress and neuroinflammation. Neurobiol Stress 2016;4:23–33. 10.1016/j.ynstr.2016.03.001.

[18] Lee YK, Menezes JS, Umesaki Y, Mazmanian SK. Proinflammatory T-cell responses to gut microbiota promote experimental autoimmune encephalomyelitis. Proc Natl Acad Sci 2011;108:4615–22. 10.1073/pnas.1000082107.

[19] Seo D, O’Donnell D, Jain N, Ulrich JD, Herz J, Li Y, et al. ApoE isoform– and microbiota-dependent progression of neurodegeneration in a mouse model of tauopathy. Science 2023;379:eadd1236. 10.1126/science.add1236.

[20] Nikolova VL, Smith MRB, Hall LJ, Cleare AJ, Stone JM, Young AH. Perturbations in Gut Microbiota Composition in Psychiatric Disorders: A Review and Meta-analysis. JAMA Psychiatry 2021;78:1343. 10.1001/jamapsychiatry.2021.2573.

[21] Sekula P, Del Greco MF, Pattaro C, Köttgen A. Mendelian Randomization as an Approach to Assess Causality Using Observational Data. J Am Soc Nephrol 2016;27:3253–65. 10.1681/ASN.2016010098.

[22] Emdin CA, Khera AV, Kathiresan S. Mendelian Randomization. JAMA 2017;318:1925. 10.1001/jama.2017.17219.

[23] Larsson SC, Butterworth AS, Burgess S. Mendelian randomization for cardiovascular diseases: principles and applications. Eur Heart J 2023;44:4913–24. 10.1093/eurheartj/ehad736.

[24] Skrivankova VW, Richmond RC, Woolf BAR, Yarmolinsky J, Davies NM, Swanson SA, et al. Strengthening the Reporting of Observational Studies in Epidemiology Using Mendelian Randomization: The STROBE-MR Statement. JAMA 2021;326:1614. 10.1001/jama.2021.18236.

[25] Orrù V, Steri M, Sidore C, Marongiu M, Serra V, Olla S, et al. Complex genetic signatures in immune cells underlie autoimmunity and inform therapy. Nat Genet 2020;52:1036–45. 10.1038/s41588-020-0684-4.

[26] Lopera-Maya EA, Kurilshikov A, Van Der Graaf A, Hu S, Andreu-Sánchez S, Chen L, et al. Effect of host genetics on the gut microbiome in 7,738 participants of the Dutch Microbiome Project. Nat Genet 2022;54:143–51. 10.1038/s41588-021-00992-y.

[27] Demontis D, Walters GB, Athanasiadis G, Walters R, Therrien K, Nielsen TT, et al. Genome-wide analyses of ADHD identify 27 risk loci, refine the genetic architecture and implicate several cognitive domains. Nat Genet 2023;55:198–208. 10.1038/s41588-022-01285-8.

[28] Howard DM, Adams MJ, Clarke T-K, Hafferty JD, Gibson J, Shirali M, et al. Genome-wide meta-analysis of depression identifies 102 independent variants and highlights the importance of the prefrontal brain regions. Nat Neurosci 2019;22:343–52. 10.1038/s41593-018-0326-7.

[29] Nievergelt CM, Maihofer AX, Klengel T, Atkinson EG, Chen C-Y, Choi KW, et al. International meta-analysis of PTSD genome-wide association studies identifies sex- and ancestry-specific genetic risk loci. Nat Commun 2019;10:4558. 10.1038/s41467-019-12576-w.

[30] Trubetskoy V, Pardiñas AF, Qi T, Panagiotaropoulou G, Awasthi S, Bigdeli TB, et al. Mapping genomic loci implicates genes and synaptic biology in schizophrenia. Nature 2022;604:502–8. 10.1038/s41586-022-04434-5.

[31] Anorexia Nervosa Genetics Initiative, Eating Disorders Working Group of the Psychiatric Genomics Consortium, Watson HJ, Yilmaz Z, Thornton LM, Hübel C, et al. Genome-wide association study identifies eight risk loci and implicates metabo-psychiatric origins for anorexia nervosa. Nat Genet 2019;51:1207–14. 10.1038/s41588-019-0439-2.

[32] Shen H-H, Zhang Y-Y, Wang X-Y, Wang C-J, Wang Y, Ye J-F, et al. Potential Causal Association between Plasma Metabolites, Immunophenotypes, and Female Reproductive Disorders: A Two-Sample Mendelian Randomization Analysis. Biomolecules 2024;14:116. 10.3390/biom14010116.

[33] Ji D, Chen W-Z, Zhang L, Zhang Z-H, Chen L-J. Gut microbiota, circulating cytokines and dementia: a Mendelian randomization study. J Neuroinflammation 2024;21:2. 10.1186/s12974-023-02999-0.

[34] The 1000 Genomes Project Consortium, Corresponding authors, Auton A, Abecasis GR, Steering committee, Altshuler DM, et al. A global reference for human genetic variation. Nature 2015;526:68–74. 10.1038/nature15393.

[35] Guo B, Wang C, Zhu Y, Liu Z, Long H, Ruan Z, et al. Causal associations of brain structure with bone mineral density: a large-scale genetic correlation study. Bone Res 2023;11:37. 10.1038/s41413-023-00270-z.

[36] Tang M, Wang T, Zhang X. A review of SNP heritability estimation methods. Brief Bioinform 2022;23:bbac067. 10.1093/bib/bbac067.

[37] Burgess S, Thompson SG, CRP CHD Genetics Collaboration. Avoiding bias from weak instruments in Mendelian randomization studies. Int J Epidemiol 2011;40:755–64. 10.1093/ije/dyr036.

[38] Hemani G, Zheng J, Elsworth B, Wade KH, Haberland V, Baird D, et al. The MR-Base platform supports systematic causal inference across the human phenome. eLife 2018;7:e34408. 10.7554/eLife.34408.

[39] Slob EAW, Burgess S. A comparison of robust Mendelian randomization methods using summary data. Genet Epidemiol 2020;44:313–29. 10.1002/gepi.22295.

[40] Bowden J, Davey Smith G, Burgess S. Mendelian randomization with invalid instruments: effect estimation and bias detection through Egger regression. Int J Epidemiol 2015;44:512–25. 10.1093/ije/dyv080.

[41] Bowden J, Davey Smith G, Haycock PC, Burgess S. Consistent Estimation in Mendelian Randomization with Some Invalid Instruments Using a Weighted Median Estimator. Genet Epidemiol 2016;40:304–14. 10.1002/gepi.21965.

[42] Glickman ME, Rao SR, Schultz MR. False discovery rate control is a recommended alternative to Bonferroni-type adjustments in health studies. J Clin Epidemiol 2014;67:850–7. 10.1016/j.jclinepi.2014.03.012.

[43] Ji D, Chen W-Z, Zhang L, Zhang Z-H, Chen L-J. Gut microbiota, circulating cytokines and dementia: a Mendelian randomization study. J Neuroinflammation 2024;21:2. 10.1186/s12974-023-02999-0.

[44] Wang Q, Dai H, Hou T, Hou Y, Wang T, Lin H, et al. Dissecting Causal Relationships Between Gut Microbiota, Blood Metabolites, and Stroke: A Mendelian Randomization Study. J Stroke 2023;25:350–60. 10.5853/jos.2023.00381.

[45] Kulinskaya E, Dollinger MB, Bjørkestøl K. On the moments of Cochran’s Q statistic under the null hypothesis, with application to the meta[analysis of risk difference. Res Synth Methods 2020;11:920–920. 10.1002/jrsm.1446.

[46] Burgess S, Bowden J, Fall T, Ingelsson E, Thompson SG. Sensitivity Analyses for Robust Causal Inference from Mendelian Randomization Analyses with Multiple Genetic Variants. Epidemiology 2017;28:30–42. 10.1097/EDE.0000000000000559.

[47] Burgess S, Thompson SG. Interpreting findings from Mendelian randomization using the MR-Egger method. Eur J Epidemiol 2017;32:377–89. 10.1007/s10654-017-0255-x.

[48] Verbanck M, Chen C-Y, Neale B, Do R. Detection of widespread horizontal pleiotropy in causal relationships inferred from Mendelian randomization between complex traits and diseases. Nat Genet 2018;50:693–8. 10.1038/s41588-018-0099-7.

[49] Hemani G, Tilling K, Davey Smith G. Orienting the causal relationship between imprecisely measured traits using GWAS summary data. PLOS Genet 2017;13:e1007081. 10.1371/journal.pgen.1007081.

[50] Hemani G, Zheng J, Elsworth B, Wade KH, Haberland V, Baird D, et al. The MR-Base platform supports systematic causal inference across the human phenome. eLife 2018;7:e34408. 10.7554/eLife.34408.

[51] Verbanck M, Chen C-Y, Neale B, Do R. Detection of widespread horizontal pleiotropy in causal relationships inferred from Mendelian randomization between complex traits and diseases. Nat Genet 2018;50:693–8. 10.1038/s41588-018-0099-7.

[52] Park JH. Potential Inflammatory Biomarker in Patients with Attention Deficit Hyperactivity Disorder. Int J Mol Sci 2022;23:13054. 10.3390/ijms232113054.

[53] Faraone SV, Asherson P, Banaschewski T, Biederman J, Buitelaar JK, Ramos-Quiroga JA, et al. Attention-deficit/hyperactivity disorder. Nat Rev Dis Primer 2015;1:15020. 10.1038/nrdp.2015.20.

[54] Tamouza R, Krishnamoorthy R, Leboyer M. Understanding the genetic contribution of the human leukocyte antigen system to common major psychiatric disorders in a world pandemic context. Brain Behav Immun 2021;91:731–9. 10.1016/j.bbi.2020.09.033.

[55] Van De Wouw M, Boehme M, Lyte JM, Wiley N, Strain C, O’Sullivan O, et al. Short[chain fatty acids: microbial metabolites that alleviate stress[induced brain–gut axis alterations. J Physiol 2018;596:4923–44. 10.1113/JP276431.

[56] Wang Y, Subudhi SK, Anders RA, Lo J, Sun Y, Blink S, et al. The role of herpesvirus entry mediator as a negative regulator of T cell–mediated responses. J Clin Invest 2005;115:711–7. 10.1172/JCI200522982.

[57] Kaech SM, Wherry EJ, Ahmed R. Effector and memory T-cell differentiation: implications for vaccine development. Nat Rev Immunol 2002;2:251–62. 10.1038/nri778.

[58] Jain A, Song R, Wakeland EK, Pasare C. T cell-intrinsic IL-1R signaling licenses effector cytokine production by memory CD4 T cells. Nat Commun 2018;9:3185. 10.1038/s41467-018-05489-7.

[59] Freeman S, Kelm S, Barber E, Crocker P. Characterization of CD33 as a new member of the sialoadhesin family of cellular interaction molecules. Blood 1995;85:2005–12. 10.1182/blood.V85.8.2005.bloodjournal8582005.

[60] Crocker PR, Paulson JC, Varki A. Siglecs and their roles in the immune system. Nat Rev Immunol 2007;7:255–66. 10.1038/nri2056.

[61] Lukens JR, Eyo UB. Microglia and Neurodevelopmental Disorders. Annu Rev Neurosci 2022;45:425–45. 10.1146/annurev-neuro-110920-023056.

[62] Mordelt A, De Witte LD. Microglia-mediated synaptic pruning as a key deficit in neurodevelopmental disorders: Hype or hope? Curr Opin Neurobiol 2023;79:102674. 10.1016/j.conb.2022.102674.

[63] Ge Y, Cheng D, Jia Q, Xiong H, Zhang J. Mechanisms Underlying the Role of Myeloid-Derived Suppressor Cells in Clinical Diseases: Good or Bad. Immune Netw 2021;21:e21. 10.4110/in.2021.21.e21.

[64] Melero-Jerez C, Ortega MC, Moliné-Velázquez V, Clemente D. Myeloid derived suppressor cells in inflammatory conditions of the central nervous system. Biochim Biophys Acta BBA - Mol Basis Dis 2016;1862:368–80. 10.1016/j.bbadis.2015.10.015.

[65] Newman TM, Shively CA, Register TC, Appt SE, Yadav H, Colwell RR, et al. Diet, obesity, and the gut microbiome as determinants modulating metabolic outcomes in a non-human primate model. Microbiome 2021;9:100. 10.1186/s40168-021-01069-y.

[66] Reichardt N, Duncan SH, Young P, Belenguer A, McWilliam Leitch C, Scott KP, et al. Phylogenetic distribution of three pathways for propionate production within the human gut microbiota. ISME J 2014;8:1323–35. 10.1038/ismej.2014.14.

[67] Notting F, Pirovano W, Sybesma W, Kort R. The butyrate-producing and spore-forming bacterial genus *Coprococcus* as a potential biomarker for neurological disorders. Gut Microbiome 2023;4:e16. 10.1017/gmb.2023.14.

[68] Shalev A, Liberzon I, Marmar C. Post-Traumatic Stress Disorder. N Engl J Med 2017;376:2459–69. 10.1056/NEJMra1612499.

[69] Girgenti MJ, Duman RS. Transcriptome Alterations in Posttraumatic Stress Disorder. Biol Psychiatry 2018;83:840–8. 10.1016/j.biopsych.2017.09.023.

[70] Antshel KM, Kaul P, Biederman J, Spencer TJ, Hier BO, Hendricks K, et al. Posttraumatic Stress Disorder in Adult Attention-Deficit/Hyperactivity Disorder: Clinical Features and Familial Transmission. J Clin Psychiatry 2013;74:e197–204. 10.4088/JCP.12m07698.

[71] Herrero ER, Slomka V, Bernaerts K, Boon N, Hernandez-Sanabria E, Passoni BB, et al. Antimicrobial effects of commensal oral species are regulated by environmental factors. J Dent 2016;47:23–33. 10.1016/j.jdent.2016.02.007.

[72] Garnett JA, Simpson PJ, Taylor J, Benjamin SV, Tagliaferri C, Cota E, et al. Structural insight into the role of Streptococcus parasanguinis Fap1 within oral biofilm formation. Biochem Biophys Res Commun 2012;417:421–6. 10.1016/j.bbrc.2011.11.131.

[73] Li S, Li N, Wang C, Zhao Y, Cao J, Li X, et al. Gut Microbiota and Immune Modulatory Properties of Human Breast Milk Streptococcus salivarius and S. parasanguinis Strains. Front Nutr 2022;9:798403. 10.3389/fnut.2022.798403.

[74] Bozzatello P, De Rosa ML, Rocca P, Bellino S. Effects of Omega 3 Fatty Acids on Main Dimensions of Psychopathology. Int J Mol Sci 2020;21:6042. 10.3390/ijms21176042.

[75] Guerreiro S, Privat A-L, Bressac L, Toulorge D. CD38 in Neurodegeneration and Neuroinflammation. Cells 2020;9:471. 10.3390/cells9020471.

[76] Zhang X. IL-10 is involved in the suppression of experimental autoimmune encephalomyelitis by CD25+CD4+ regulatory T cells. Int Immunol 2004;16:249–56. 10.1093/intimm/dxh029.

[77] Thornton AM, Shevach EM. CD4+CD25+ Immunoregulatory T Cells Suppress Polyclonal T Cell Activation In Vitro by Inhibiting Interleukin 2 Production. J Exp Med 1998;188:287–96. 10.1084/jem.188.2.287.

[78] Michopoulos V, Powers A, Gillespie CF, Ressler KJ, Jovanovic T. Inflammation in Fear- and Anxiety-Based Disorders: PTSD, GAD, and Beyond. Neuropsychopharmacology 2017;42:254–70. 10.1038/npp.2016.146.

[79] Kahn RS, Sommer IE, Murray RM, Meyer-Lindenberg A, Weinberger DR, Cannon TD, et al. Schizophrenia. Nat Rev Dis Primer 2015;1:15067. 10.1038/nrdp.2015.67.

[80] Liu JCW, Gorbovskaya I, Hahn MK, Müller DJ. The Gut Microbiome in Schizophrenia and the Potential Benefits of Prebiotic and Probiotic Treatment. Nutrients 2021;13:1152. 10.3390/nu13041152.

[81] Jin D, Liu H-X, Hirai H, Torashima T, Nagai T, Lopatina O, et al. CD38 is critical for social behaviour by regulating oxytocin secretion. Nature 2007;446:41–5. 10.1038/nature05526.

[82] Steiner J, Jacobs R, Panteli B, Brauner M, Schiltz K, Bahn S, et al. Acute schizophrenia is accompanied by reduced T cell and increased B cell immunity. Eur Arch Psychiatry Clin Neurosci 2010;260:509–18. 10.1007/s00406-010-0098-x.

[83] Melbourne JK, Rosen C, Chase KA, Feiner B, Sharma RP. Monocyte Transcriptional Profiling Highlights a Shift in Immune Signatures Over the Course of Illness in Schizophrenia. Front Psychiatry 2021;12:649494. 10.3389/fpsyt.2021.649494.

[84] Schramm E, Klein DN, Elsaesser M, Furukawa TA, Domschke K. Review of dysthymia and persistent depressive disorder: history, correlates, and clinical implications. Lancet Psychiatry 2020;7:801–12. 10.1016/S2215-0366(20)30099-7.

[85] Gao X, Tang Y, Kong L, Fan Y, Wang C, Wang R. Treg cell: Critical role of regulatory T-cells in depression. Pharmacol Res 2023;195:106893. 10.1016/j.phrs.2023.106893.

[86] Schiepers OJG, Wichers MC, Maes M. Cytokines and major depression. Prog Neuropsychopharmacol Biol Psychiatry 2005;29:201–17. 10.1016/j.pnpbp.2004.11.003.

[87] Schlatter J, Ortuño F, Cervera-Enguix S. Lymphocyte subsets and lymphokine production in patients with melancholic versus nonmelancholic depression. Psychiatry Res 2004;128:259–65. 10.1016/j.psychres.2004.06.004.

[88] Gao X, Tang Y, Kong L, Fan Y, Wang C, Wang R. Treg cell: Critical role of regulatory T-cells in depression. Pharmacol Res 2023;195:106893. 10.1016/j.phrs.2023.106893.

[89] Grosse L, Carvalho LA, Birkenhager TK, Hoogendijk WJ, Kushner SA, Drexhage HA, et al. Circulating cytotoxic T cells and natural killer cells as potential predictors for antidepressant response in melancholic depression. Restoration of T regulatory cell populations after antidepressant therapy. Psychopharmacology (Berl) 2016;233:1679–88. 10.1007/s00213-015-3943-9.

[90] Geng J, Liu C, Xu J, Wang X, Li X. Potential relationship between Tourette syndrome and gut microbiome. J Pediatr (Rio J) 2023;99:11–6. 10.1016/j.jped.2022.06.002.

[91] Li H, Wang Y, Zhao C, Liu J, Zhang L, Li A. Fecal transplantation can alleviate tic severity in a Tourette syndrome mouse model by modulating intestinal flora and promoting serotonin secretion. Chin Med J (Engl) 2022;135:707–13. 10.1097/CM9.0000000000001885.

[92] Martinović A, Cocuzzi R, Arioli S, Mora D. Streptococcus thermophilus: To Survive, or Not to Survive the Gastrointestinal Tract, That Is the Question! Nutrients 2020;12:2175. 10.3390/nu12082175.

[93] Khalifa N, Von Knorring A-L. Psychopathology in a Swedish Population of School Children With Tic Disorders. J Am Acad Child Adolesc Psychiatry 2006;45:1346–53. 10.1097/01.chi.0000251210.98749.83.

[94] Freeman RD, Tourette Syndrome International Database Consortium. Tic disorders and ADHD: answers from a world-wide clinical dataset on Tourette syndrome. Eur Child Adolesc Psychiatry 2007;16:15–23. 10.1007/s00787-007-1003-7.

[95] Wang Y, Xu H, Jing M, Hu X, Wang J, Hua Y. Gut Microbiome Composition Abnormalities Determined Using High-Throughput Sequencing in Children With Tic Disorder. Front Pediatr 2022;10:831944. 10.3389/fped.2022.831944.

[96] Fongang B, Satizabal C, Kautz TF, Wadop YN, Muhammad JAS, Vasquez E, et al. Cerebral small vessel disease burden is associated with decreased abundance of gut Barnesiella intestinihominis bacterium in the Framingham Heart Study. Sci Rep 2023;13:13622. 10.1038/s41598-023-40872-5.

[97] Meyer K, Lulla A, Debroy K, Shikany JM, Yaffe K, Meirelles O, et al. Association of the Gut Microbiota With Cognitive Function in Midlife. JAMA Netw Open 2022;5:e2143941. 10.1001/jamanetworkopen.2021.43941.

