## Supplementary material for "Causal effect of the gut microbiota on the risk of psychiatric disorders and the mediating role of immunophenotypes": STROBE-MR checklist

**STROBE-MR checklist of recommended items to address in reports of Mendelian randomization studies**^1,2^

| **Item No.** | **Section** | **Checklist item** | **Page No.** | **Relevant text from manuscript** |
| --- | --- | --- | --- | --- |
| **1** | **Title and Abstract** | Indicate Mendelian randomization (MR) as the study’s design in the title and/or the abstract if that is a main purpose of the study | 4 | “We utilized a bidirectional Mendelian randomization (MR) study ...” |
|  | **INTRODUCTION** |  |  |  |
| **2** | **Background** | Explain the scientific background and rationale for the reported study. What is the exposure? Is a potential causal relationship between exposure and outcome plausible? Justify why MR is a helpful method to address the study question | 6 | “Mendelian randomization is an analytical approach that utilizes single nucleotide polymorphisms (SNPs) as an instrumental variable (IV) to investigate the causal relationship between an exposure or a risk factor and a clinically relevant outcome while minimizing the risk of bias and reverse causation” |
| **3** | **Objectives** | State specific objectives clearly, including pre-specified causal hypotheses (if any). State that MR is a method that, under specific assumptions, intends to estimate causal effects | 6 | “In this study, we employed a systematic bidirectional two-sample MR design to comprehensively investigate the causal effects among the gut microbiome, immunophenotypes, and various psychiatric disorders.” |
|  | **METHODS** |  |  |  |
| **4** | **Study design and data sources** | Present key elements of the study design early in the article. Consider including a table listing sources of data for all phases of the study. For each data source contributing to the analysis, describe the following: |  |  |
|  | a) | Setting: Describe the study design and the underlying population, if possible. Describe the setting, locations, and relevant dates, including periods of recruitment, exposure, follow-up, and data collection, when available. | 7 | “The study analyzed a total of ...” |
|  | b) | Participants: Give the eligibility criteria, and the sources and methods of selection of participants. Report the sample size, and whether any power or sample size calculations were carried out prior to the main analysis | 7 | “3,757 Sardinians ... 7,738 European individuals ... The characteristics of the selected GWAS data are listed in Table S1.” |
|  | c) | Describe measurement, quality control and selection of genetic variants | 7-8 | “First, SNPs significantly associated with the immunophenotypes …” |
|  | d) | For each exposure, outcome, and other relevant variables, describe methods of assessment and diagnostic criteria for diseases | 7 | “All cohorts were case-control studies, and cases met the criteria defined in the International Statistical Classification of Diseases and Related Health Problems-10th Revision (ICD-10) diagnosis code or medication prescription.” |
|  | e) | Provide details of ethics committee approval and participant informed consent, if relevant | 22 | “The present study is a secondary analysis of publicly available data. Ethical approval was granted for each of the original GWAS. Furthermore, no individual-level data were used in this study. Consequently, no new ethical review board approval was required.” |
| **5** | **Assumptions** | Explicitly state the three core IV assumptions for the main analysis (relevance, independence and exclusion restriction) as well assumptions for any additional or sensitivity analysis | 6-7 | “To determine the unbiased causal effects of exposure on outcomes, MR analysis must satisfy three fundamental assumptions: ...” |
| **6** | **Statistical methods: main analysis** | Describe statistical methods and statistics used |  |  |
|  | a) | Describe how quantitative variables were handled in the analyses (i.e., scale, units, model) | 10 | “Effect estimates are reported as beta values (ß) for the continuous outcome and ORs for the binary outcome.” |
|  | b) | Describe how genetic variants were handled in the analyses and, if applicable, how their weights were selected | 8 | “The proportion of variance in the explained phenotype (R2) was calculated to indicate the power of MR studies. The F-statistic of each IV was calculated using the formula [R2/(1-R2)] × [(N-K-1)/K].” |
|  | c) | Describe the MR estimator (e.g. two-stage least squares, Wald ratio) and related statistics. Detail the included covariates and, in case of two-sample MR, whether the same covariate set was used for adjustment in the two samples | 8 | “Two-sample MR analysis was conducted using the multiplicative random effects inverse variance weighted approach to ...” |
|  | d) | Explain how missing data were addressed | 8 | “To compensate for missing SNPs, those with strong linkage disequilibrium (r2 > 0.8) were utilized.” |
|  | e) | If applicable, indicate how multiple testing was addressed | 8 | “The false discovery rate (FDR) was utilized to adjust for multiple tests, and a significant causal relationship was established using an FDR-adjusted threshold of P < 0.05. A P-value of less than 0.05, but above the FDR threshold, was considered indicative of a nominal association.” |
| **7** | **Assessment of assumptions** | Describe any methods or prior knowledge used to assess the assumptions or justify their validity | 8 | “To evaluate the robustness of the primary estimates, the MR-Egger and Weighted Median methods were employed to corroborate the IVW results.” |
| **8** | **Sensitivity analyses and additional analyses** | Describe any sensitivity analyses or additional analyses performed (e.g. comparison of effect estimates from different approaches, independent replication, bias analytic techniques, validation of instruments, simulations) | 9 | “Cochran’s Q statistic was used to assess the heterogeneity across ...” |
| **9** | **Software and pre-registration** |  |  |  |
|  | a) | Name statistical software and package(s), including version and settings used | 9 | “All analyses were conducted using R statistical software ...” |
|  | b) | State whether the study protocol and details were pre-registered (as well as when and where) | NA | NA |
|  | **RESULTS** |  |  |  |
| **10** | **Descriptive data** |  |  |  |
|  | a) | Report the numbers of individuals at each stage of included studies and reasons for exclusion. Consider use of a flow diagram | NA | NA |
|  | b) | Report summary statistics for phenotypic exposure(s), outcome(s), and other relevant variables (e.g. means, SDs, proportions) | 7 | “The characteristics of the selected GWAS data are listed in Table S1.” |
|  | c) | If the data sources include meta-analyses of previous studies, provide the assessments of heterogeneity across these studies | NA | NA |
|  | d) | For two-sample MR:  i.  Provide justification of the similarity of the genetic variant-exposure associations between the exposure and outcome samples  ii.  Provide information on the number of individuals who overlap between the exposure and outcome studies | 7-8 | “There was no significant overlap between the GWAS datasets. … First, SNPs significantly associated with the immunophenotypes …” |
| **11** | **Main results** |  |  |  |
|  | a) | Report the associations between genetic variant and exposure, and between genetic variant and outcome, preferably on an interpretable scale | 10 | “Further details on the specific genetic instrumental variables used to infer causal effects are provided in Table S2” |
|  | b) | Report MR estimates of the relationship between exposure and outcome, and the measures of uncertainty from the MR analysis, on an interpretable scale, such as odds ratio or relative risk per SD difference | 10-14 | “We identified 54 potential causal associations between...” |
|  | c) | If relevant, consider translating estimates of relative risk into absolute risk for a meaningful time period | NA | NA |
|  | d) | Consider plots to visualize results (e.g. forest plot, scatterplot of associations between genetic variants and outcome versus between genetic variants and exposure) |  | Figure 2 & Figure 3 |
| **12** | **Assessment of assumptions** |  |  |  |
|  | a) | Report the assessment of the validity of the assumptions | 15 | “No evidence of horizontal pleiotropy was identified …” |
|  | b) | Report any additional statistics (e.g., assessments of heterogeneity across genetic variants, such as *I^2^*, Q statistic or E-value) | 15 | “The Cochran’s Q test indicated ...” |
| **13** | **Sensitivity analyses and additional analyses** |  |  |  |
|  | a) | Report any sensitivity analyses to assess the robustness of the main results to violations of the assumptions | 15 | “No evidence of horizontal pleiotropy was identified …” |
|  | b) | Report results from other sensitivity analyses or additional analyses | 10-15 | “However, in leave-one-out analyses ...” |
|  | c) | Report any assessment of direction of causal relationship (e.g., bidirectional MR) | 15 | “According to the backward analysis …” |
|  | d) | When relevant, report and compare with estimates from non-MR analyses | NA | NA |
|  | e) | Consider additional plots to visualize results (e.g., leave-one-out analyses) | NA | NA |
|  | **DISCUSSION** |  |  |  |
| **14** | **Key results** | Summarize key results with reference to study objectives | 15 | “This study presents, for the first time, multiple novel potential associations between...” |
| **15** | **Limitations** | Discuss limitations of the study, taking into account the validity of the IV assumptions, other sources of potential bias, and imprecision. Discuss both direction and magnitude of any potential bias and any efforts to address them | 20 | “There are several limitations to consider in this study ...” |
| **16** | **Interpretation** |  |  |  |
|  | a) | Meaning: Give a cautious overall interpretation of results in the context of their limitations and in comparison with other studies | 15-20 | 2nd-8th paragraph in the Discussion |
|  | b) | Mechanism: Discuss underlying biological mechanisms that could drive a potential causal relationship between the investigated exposure and the outcome, and whether the gene-environment equivalence assumption is reasonable. Use causal language carefully, clarifying that IV estimates may provide causal effects only under certain assumptions | 15-20 | 2nd-8th paragraph in the Discussion |
|  | c) | Clinical relevance: Discuss whether the results have clinical or public policy relevance, and to what extent they inform effect sizes of possible interventions | 15-20 | 2nd-8th paragraph in the Discussion |
| **17** | **Generalizability** | Discuss the generalizability of the study results (a) to other populations, (b) across other exposure periods/timings, and (c) across other levels of exposure | 20 | “The current GWAS database constraints hinder the analysis of data from other ethnic groups, and caution is advised when interpreting our results in different populations.” |
|  | **OTHER INFORMATION** |  |  |  |
| **18** | **Funding** | Describe sources of funding and the role of funders in the present study and, if applicable, sources of funding for the databases and original study or studies on which the present study is based | 22 | “This study was supported by...” |
| **19** | **Data and data sharing** | Provide the data used to perform all analyses or report where and how the data can be accessed, and reference these sources in the article. Provide the statistical code needed to reproduce the results in the article, or report whether the code is publicly accessible and if so, where | 21 | “The datasets analyzed in this study are publicly available summary statistics. The data can be obtained through cited papers and websites.” |
| **20** | **Conflicts of Interest** | All authors should declare all potential conflicts of interest | 21 | “The authors declare that they have no competing interests.” |

This checklist is copyrighted by the Equator Network under the Creative Commons Attribution 3.0 Unported (CC BY 3.0) license.

1.	Skrivankova VW, Richmond RC, Woolf BAR, Yarmolinsky J, Davies NM, Swanson SA, et al. Strengthening the Reporting of Observational Studies in Epidemiology using Mendelian Randomization (STROBE-MR) Statement. JAMA. 2021;under review.

2.	Skrivankova VW, Richmond RC, Woolf BAR, Davies NM, Swanson SA, VanderWeele TJ, et al. Strengthening the Reporting of Observational Studies in Epidemiology using Mendelian Randomisation (STROBE-MR): Explanation and Elaboration. BMJ. 2021;375:n2233.
